# Social-semantic knowledge in frontotemporal dementia and after anterior temporal lobe resection

**DOI:** 10.1101/2024.04.11.24305610

**Authors:** Matthew A. Rouse, Ajay D. Halai, Siddharth Ramanan, Timothy T. Rogers, Peter Garrard, Karalyn Patterson, James B. Rowe, Matthew A. Lambon Ralph

**Affiliations:** MRC Cognition and Brain Sciences Unit, University of Cambridge, Cambridge, CB2 7EF, UK; Department of Psychology, University of Wisconsin-Madison, Madison, WI 53706, USA; Molecular and Clinical Sciences Research Institute, St George’s, University of London, London, SW17 0RE, UK; Department of Clinical Neurosciences, University of Cambridge, Cambridge, CB2 0SZ, UK; Cambridge University Hospitals NHS Foundation Trust, Cambridge, CB2 0SZ, UK

**Keywords:** behavioural-variant frontotemporal dementia, principal component analysis, semantic dementia, social concept, temporal lobe epilepsy, transdiagnostic

## Abstract

Degraded semantic memory is a prominent feature of frontotemporal dementia (FTD). It is classically associated with semantic dementia and anterior temporal lobe (ATL) atrophy, but semantic knowledge can also be compromised in behavioural-variant FTD (bvFTD). Motivated by understanding behavioural change in FTD, recent research has focused selectively on social-semantic knowledge, with proposals that the right ATL is specialised for social concepts. Previous studies have assessed very different types of social concepts and have not compared performance to that on matched non-social concepts. Consequently, it remains unclear to what extent various social concepts are (i) concurrently impaired in FTD, (ii) distinct from general semantic memory and (iii) differentially supported by the left and right ATL. This study assessed multiple aspects of social-semantic knowledge and general conceptual knowledge across cohorts with ATL-damage arising from either neurodegeneration or resection. We assembled a test battery measuring knowledge of multiple types of social concept. Performance was compared to non-social general conceptual knowledge, measured using the Cambridge Semantic Memory Test Battery and other matched non-social-semantic tests. Our transdiagnostic approach included bvFTD, semantic dementia and “mixed” intermediate cases to capture the FTD clinical spectrum, as well as age-matched healthy controls. People with unilateral left or right ATL resection for temporal lobe epilepsy (TLE) were also recruited to assess how selective damage to the left or right ATL impacts social- and non-social-semantic knowledge. Social- and non-social-semantic deficits were severe and highly correlated in FTD. Much milder impairments were found after unilateral ATL resection, with no left vs. right differences in social-semantic knowledge or general semantic processing, and with only naming showing a greater deficit following left vs. right damage. A principal component analysis of all behavioural measures in the FTD cohort extracted three components, interpreted as capturing: (1) FTD severity, (2) semantic memory and (3) executive function. Social and non-social measures both loaded heavily on the same semantic memory component, and scores on this factor were uniquely associated with bilateral ATL grey matter volume but not with the degree of ATL asymmetry. Together, these findings demonstrate that both social- and non-social-semantic knowledge degrade in FTD (semantic dementia and bvFTD) following bilateral ATL atrophy. We propose that social-semantic knowledge is part of a broader conceptual system underpinned by a bilaterally-implemented, functionally-unitary semantic hub in the ATLs. Our results also highlight the value of a transdiagnostic approach for investigating the neuroanatomical underpinnings of cognitive deficits in FTD.

## Introduction

Degraded semantic memory is a prominent feature of frontotemporal dementia (FTD). It is classically associated with semantic dementia (SD; also called semantic-variant primary progressive aphasia (svPPA)) and atrophy in the anterior temporal lobes (ATLs),^1–6^ but is also often a feature in behavioural-variant FTD (bvFTD).^7–9^ Motivated by the behavioural changes that are commonly observed in FTD, a line of recent research has focussed on a specific aspect of the conceptual system, *social-semantic knowledge*, and the potentially pivotal importance of the right ATL.^10–16^ The degree to which this type of knowledge is (i) impaired in FTD, (ii) concurrently impaired across different types of social concept, (iii) distinct from general semantic memory and (iv) supported by the left and right ATLs is unclear. To address this gap in clinical knowledge, and to better understand whether social- and non-social-semantic knowledge are distinct domains with different neuroanatomical underpinnings, we assembled a novel “broadband” battery spanning the many different types of social concept that have often been assessed only singly in past studies. An FTD cohort was recruited, including bvFTD, SD (including svPPA (commonly L>R ATL atrophy) and R>L ATL “right” SD) and “mixed” intermediate cases to ensure full coverage of the FTD clinical space and the underlying variations in atrophy across the associated frontotemporal neuroanatomy. To provide important convergent data on the function of the left and right ATL, people with left or right unilateral ATL resection for temporal lobe epilepsy (TLE) also took part. Social-semantic performance was compared with general conceptual knowledge, assessed using the Cambridge Semantic Memory Test Battery^17,18^ and other matched non-social semantic tasks. Thus, for the first time, we were able to test multiple aspects of social-semantic knowledge in parallel and compare this to general conceptual knowledge in FTD and after ATL resection. For the purposes of the current study, we consider “semantic” and “conceptual” knowledge as synonymous, although we note that some disciplines draw distinctions between the two terms.^19^

Separate investigations in clinical and cognitive neuroscience have highlighted roles for the ATLs in semantic memory^20–22^ and/or social cognition.^11,23–25^ People with SD experience a degradation of semantic memory following bilateral ATL atrophy^2–4,6,18^ and also display behavioural changes.^26–28^ In their severest form, these semantic and behavioural changes are reminiscent of the classic Klüver and Bucy studies which found concurrent multimodal associative agnosia and chronic behaviour change following bilateral (but not unilateral) ATL ablation in macaques.^29^ Provided appropriate techniques which maximise ventral ATL signal are used,^30^ contemporary fMRI studies have detected bilateral ventrolateral ATL activation when healthy participants engage in semantic processing for all types of concepts^20,31^ including social concepts^32,33^ and for other aspects of social cognition such as theory of mind.^23^ One explanation for a shared contribution to semantic and social processing is that the ATLs store social knowledge as part of semantic memory more generally.^10,13^ A challenge of the current literature in providing clear answers to these questions is that there is no general consensus on what makes a concept “social”^13,34,35^ and thus it is unclear what types of concept are critical to social behaviours. Past investigations have each tended to focus on one type of “social” concept, which collectively span a very diverse range of concrete-to-abstract concepts, including people,^14^ social behaviours^12,16,36^ and emotions.^37–39^

The neuroanatomical basis of social-semantic knowledge is also a subject of current debate.^13,35,40^ According to one hypothesis, the right ATL is specialised for social-semantic knowledge, whereas the left ATL supports verbal semantic knowledge.^14,16^ This dichotomy is largely based on the clinical observations of R > L SD patients (also sometimes known as right-temporal variant FTD), who often have prosopagnosia in the very earliest stages (typically before most patients present to clinic)^1,41^ followed by the emergence of behavioural changes and a generalised semantic impairment.^14,42–45^ This clinical evidence accords with more formal research showing that rightward-biased ATL atrophy/hypometabolism is associated with deficits in person knowledge,^14^ social conceptual knowledge,^16^ emotion recognition^46^ and theory of mind.^47,48^ Direct comparisons between left versus right ATL atrophy in SD are not straightforward, however: even if asymmetrical, the pathology is always bilateral, making it hard to unpick the relative contributions of each side.^1,49,50^ Indeed, L > R SD patients can also develop behavioural impairment. Furthermore, R > L patients typically present to clinic later than L > R patients and, consequently, often have more severe temporal lobe atrophy^1,42^ and increased atrophy in prefrontal regions important for social behaviour.^51,52^

Recently, social-semantic knowledge has been integrated within the hub-and-spokes model of semantic memory.^10,12,13^ According to this framework, the bilateral ATLs underpin a transmodal, transtemporal hub for all concepts, which supports semantic representation through interaction with modality-specific cortical “spokes”.^21,22^ Accordingly, social-semantic knowledge is not a ‘special’ type of knowledge with a distinct neural architecture, but is part of a broader conceptual system supported by the same bilateral ATL hub as non-social concepts.^10,12,13^ A key advantage of this framework is that it not only explains the FTD data but assimilates findings from other patient groups and healthy participants. First, a recent study identified that social-semantic impairments in FTD were associated with *<u>bilateral</u>* ATL atrophy.^12^ Second, there are indications that selective right ATL damage may not cause a selective impairment for social concepts or behaviour change.^49,53^ Rather, unilateral ATL damage yields a mild general semantic impairment,^54,55^ with similarly subtle deficits in person knowledge and emotion recognition after left or right resection.^49,53^ Third, there is no strong evidence for a left/right difference in social conceptual processing from studies in healthy participants. Whereas block-design fMRI studies have found right superior ATL activation for social concepts,^15,56^ a separate study found that social-stimuli activated the *left* superior ATL.^57^ Contemporary distortion-corrected or -reducing fMRI studies have found bilateral ventrolateral ATL activation for both social and non-social concepts (although with some selectivity for social concepts in the bilateral superior ATL).^32,33,58^ A transcranial magnetic stimulation (TMS) study found that left *or* right superior ATL stimulation caused a cognitively and anatomically-selective disruption to social conceptual decision making, although stimulation to the right superior ATL yielded a relatively greater impairment to social than to non-social processing.^59^

In this study, we investigated social-semantic knowledge in two clinical groups associated with ATL damage – neurodegenerative FTD and surgical ATL resection. Our study was designed to overcome two methodological issues from previous studies which would help to determine the neural basis of social-semantic knowledge. First, we took an ‘inclusive’ approach with a broad range of social concepts, as well as carefully matched general (i.e., non-social) semantic tasks. This is critical to clarify whether social-semantic deficits are (a) selective to a specific type of social concept, (b) reflective of a domain-specific social-semantic impairment, or (c) part of a broader domain-general conceptual degradation. Second, comparisons of diagnostic groups defined categorically were supplemented by multivariate analytics that accommodate for the cognitive and neuroanatomical systematic variation in FTD. By positioning individuals along graded dimensions, it is possible simultaneously to model the contribution of total ATL volume, ATL laterality, and volume loss in other brain regions to social-semantic knowledge. In contrast to FTD, people with unilateral ATL resection provide a more selective lesion model of the left vs. right ATL. Inclusion of these participants thus provided important and novel cross-aetiological data on the impact of (i) unilateral vs. bilateral and (ii) left vs. right ATL damage on social-semantic knowledge. In summary, this study had broad coverage and high systematicity with respect to the materials, participants and analysis.

## Materials and methods

### Participants

Forty-eight people with FTD were recruited from dementia clinics in Addenbrooke’s Hospital, Cambridge (*N* = 40), St George’s Hospital, London (*N* = 4) and John Radcliffe Hospital, Oxford (*N* = 4). Twenty-six patients had a primary diagnosis of bvFTD^60^ and 22 met diagnostic criteria for SD^5^. Eighteen people who had undergone unilateral ATL resection for TLE (left = 11, right = 7) were recruited from Salford Royal Hospital, Manchester and the Walton Centre, Liverpool. All the TLE cases were left language dominant based on Wada testing and at least 12 months post-surgery. Nineteen healthy controls (age-matched to the FTD cohort) were recruited from the MRC Cognition and Brain Sciences Unit, University of Cambridge. All participants provided written informed consent obtained according to the Declaration of Helsinki. If participants lacked capacity to consent, their next of kin was consulted using the ‘personal consultee’ process as established by UK law. Demographic and clinical information is reported in Table 1.

**Table 1.** Demographic and clinical information for each group.

|  | Control | bvFTD | SD | Left<br>TLE | Right<br>TLE | Group difference | Post-hoc |
| --- | --- | --- | --- | --- | --- | --- | --- |
| N | 19 | 26 | 22 | 11 | 7 | - | - |
| Sex (M:F) | 9:10 | 18:8 | 8:14 | 6:5 | 3:4 | $\chi^2 = 5.67^a$, ns | - |
| Age (years) | 64.4 (6.7) | 64.3 (9.1) | 66.1 (6.8) | 46.8 (11.4) | 53.1 (9.7) | **H(4) = 28.4<sup>b</sup>, P < 0.0001** | L < C, bvFTD, SD; R < SD |
| Education (years) | 15.6 (3.4) | 11.5 (1.9) | 13.7 (2.9) | 13.4 (2.8) | 13.7 (2.1) | **H(4) = 20.3<sup>b</sup>, P < 0.001** | bvFTD < C |
| Years since symptom onset | - | 6.1 (3.5) | 5.8 (3.3) | - | - | $W_s = 286^c$, ns | - |
| Years since diagnosis | - | 1.7 (1.6) | 2.3 (1.8) | - | - | $W_s = 223^c$, ns | - |
| Years since resection | - | - | - | 11.0 (3.7) | 15.9 (2.4) | **t = 3.35<sup>d</sup>, P < 0.01** | - |
| Number of anti-epileptic drugs | - | - | - | 2.2 (1.3) | 1.6 (1.3) | t = 0.99<sup>d</sup>, ns | - |

<sup>a</sup>Chi-square test, <sup>b</sup>Kruskal-Wallis test; <sup>c</sup>Wilcoxon rank-sum test; <sup>d</sup>Independent t-test
Mean and standard deviations are reported for each group. Significant p-values are highlighted in bold.
bvFTD = behavioural-variant frontotemporal dementia, C = control, L=left TLE, ns = not significant, R=right TLE, SD = semantic dementia, TLE = temporal lobe epilepsy

### Neuropsychology

#### General semantic memory and background neuropsychology

General semantic memory was assessed using a battery of tasks across a range of verbal and non-verbal modalities. Tests included the modified picture version of the Camel and Cactus Test (CCT) and naming task from the Cambridge Semantic Memory Test Battery,^17,18,61,62^ a synonym judgement task,^61,63^ and the 30-item Boston Naming Test.^64,65^

Global cognition was assessed using the Addenbrooke’s Cognitive Examination-Revised (ACE-R), which provides a total score as well as five subscales: Attention and Orientation, Memory, Language, Fluency and Visuospatial Function.^66^ The Brixton Spatial Anticipation Test^67^ and Raven’s Coloured Progressive Matrices Set B^68^ were used to assess executive function. Full details of each task are reported in the Supplementary material.

#### Social-semantic battery

As mentioned in the Introduction, the term ‘social’ has been used widely and variably to describe multiple different types of task/stimuli (e.g., person knowledge,^14^ emotional cognition^37^ and abstract social behaviours).^16^ Rather than imposing an *a priori* single definition of ‘social concept’, we assembled multiple types of task/stimuli into one test battery. A literature review was conducted to identify the different types of concepts that have been deemed ‘social’ in past studies, meaning that the battery included a diverse set of tasks (described in detail below). Where possible, ‘non-social’ comparator tasks were included for each social-semantic task, matched for key variables such as lexical frequency, imageability and specificity.

##### Person knowledge

Person knowledge was assessed using face-to-name and face-to-profession matching tasks.^49^ Participants also completed a landmark-to-name matching task,^49^ which was included to assess non-social yet specific-level, or ‘unique entity’ concepts.^69,70^ Perceptual face matching was assessed using a 22-item task that required matching photographs of faces with different photos of the same person.^49,71^ Half of the trials used famous faces as items, whereas the other half used unfamiliar faces.

##### Abstract social concepts

Comprehension of abstract social concepts was assessed using a verbal abstract social synonym judgement task that has been utilised previously not only in patient assessment but also in studies of healthy participants.^15,16,32,33,36,59^ Participants also completed an abstract non-social synonym judgement task with items matched to the social concepts for lexical frequency, imageability and semantic diversity.^32,72^

##### Social word-picture matching

Participants completed a four-alternative forced-choice (4AFC) word-picture matching task. In each of the 35 trials, the presented word denoted a type of person with a characteristic age and/or gender (e.g. ‘infant’, ‘woman,’ ‘uncle’, etc.) and options were colour photographs of individuals. The participants also completed a non-social 4AFC word-picture matching task with words denoting manmade objects that were individually matched to the social items for lexical frequency.

##### Emotion knowledge

Two tests of emotion knowledge were employed: a ‘basic emotion’ recognition task using 19 stimuli from the Face and Gesture Recognition Network Database^73^ and a 23-item ‘complex emotion’ recognition task using more nuanced words such as embarrassment and jealousy, drawn from the Cambridge Mind Reading Face Battery (children’s version).^74^ In each task, participants were shown dynamic video clips of a person displaying an emotion and were instructed to point to the word best matching the emotion, from four response options.

##### Social norms knowledge

The Social Norms Questionnaire (SNQ) includes 22 items describing a behaviour. Participants answer whether it would be socially appropriate to perform each behaviour in the presence of a stranger or acquaintance (i.e. not a close friend or family member). The wording of some items was modified to UK-English, with permission from Dr. Katherine Rankin, developer of the questionnaire (Supplementary material).

##### Sarcasm detection

Participants completed the Social Inference-Minimal Test from the Awareness of Social Inference Test (TASIT-SM) which assesses the ability to detect sarcasm from paralinguistic cues.^75^

#### Statistical analysis

Group comparisons were assessed using R studio version 4.0.3.^76^ Normality of data and equality of variance were assessed using Shapiro-Wilk tests and Levene’s tests. Where data were normally distributed, one-way ANOVAs and post-hoc Tukey’s range tests were conducted if there was equality of variance across groups, whereas Welch ANOVAs and post-hoc Games Howell tests were conducted if variances were unequal. Where data were not normally distributed, Krukal-Wallis tests and post-hoc Dunn’s tests were conducted. A threshold of *P* < 0.05 was used to determine statistical significance.

### Single-case explorations

Single-case level analyses were conducted to determine whether there were any individual patients with selective deficits for social or non-social concepts. Selective deficits were calculated based on performance on the abstract social- and non-social synonym judgement tasks, and thus was a direct replication of the analysis strategy used in a previous study.^36^ Revised Standardised Difference Tests were conducted for each patient, to determine whether they met criteria for a *strong dissociation* (impaired on tasks X and Y and the difference in scores between the two tasks is significantly larger than in controls) or *classical dissociation* (impaired on task X but not task Y (or vice versa) and the difference in scores on the two tasks is significantly larger than in controls).^77^ For each test (*N* = 56), *P* < 0.0009 was the threshold used to determine statistical significance, adjusted for multiple comparisons using the Bonferroni correction.

### Structural MRI

#### MRI acquisition and preprocessing

Sixty-nine participants had a T1-weighted 3T structural MRI scan on a Siemens PRISMA at the University of Cambridge, Wolfson Brain Imaging Centre (bvFTD = 14, SD = 6, control = 19) or the University of Cambridge MRC Cognition and Brain Sciences Unit (bvFTD = 1, SD = 13, control = 16). Sixteen ATL-resected participants (left TLE = 9, right TLE = 7) and a separate cohort of 20 age-matched controls had a T1-weighed 3T structural MRI scan on a Philips Achieva scanner at the Manchester Clinical Research Facility, University of Manchester. Raw MRI data were converted to the Brain Imaging Dataset format^78^ and pre-processed using the Computational Anatomy Toolbox version 12 in SPM 12.^79^ Images were segmented into grey matter, white matter and CSF, and modulated and normalised to MNI space using geodesic shooting.^80^ Normalised grey matter images were spatially smoothed using a Gaussian kernel with 10mm FWHM.

#### Grey matter differences between groups

Voxel-based morphometry (VBM) was conducted to explore grey matter differences between groups. Separate general linear models were built with age, intracranial volume (ICV) and scanner site as covariates, and groups compared using independent t-tests. An explicit objective average-based mask was used, which is recommended for VBM of severely atrophic brains.^81^ Significant clusters were extracted using a cluster-level threshold of *Q* < 0.05, based on an initial voxel-level threshold of *P* < 0.001. Results were visualised using the xjView toolbox (https://www.alivelearn.net/xjview) and brain regions were labelled using the automated labelling atlas 3.^82^

#### Grey matter indices in frontotemporal regions of interest

For each participant, grey matter volume indices were calculated in two regions atrophied in FTD – the ATL and orbitofrontal cortex (OFC). The ATL masks were derived from a previous meta-analysis^58^ and the OFC masks were derived from the Harvard-Oxford Cortical Atlas (Supplementary Fig. 1). Grey matter intensity values for each region of interest (ROI) were extracted, and linear regression models fitted using the control data with each ROI as the dependent variable and age, ICV and scanner site as regressors. Each patient’s data were plugged into the model, and the residuals used to calculate two indices per brain region: *magnitude* (left + right residual) and *asymmetry* (left - right residual).

### Extracting neuropsychological components

A standard principal component analysis (PCA) with varimax-rotation was conducted on all neuropsychological tasks in the FTD cohort to extract the underlying dimensions of variation in the data. Raw scores were converted to percentages and missing data were imputed using probabilistic principal component analysis (PPCA).^83,84^ As PPCA requires the number of extracted principal components to be pre-specified, k-fold cross validation was used to determine the optimum number of components for missing data imputation.^85^ A three-component solution had the lowest root means squared error, and thus PPCA was conducted with three components. Participants were scored at chance level on tasks they were too impaired to complete. A PCA was then conducted on the full FTD sample (*N* = 48) with missing data imputed. The number of principal components was determined using the elbow method on the scree plot of eigenvalues^86^ and factor scores were calculated using the regression method. Sampling adequacy and suitability of the data for PCA were assessed using the Keiser-Meyer-Olkin (KMO) test and Bartlett’s test of sphericity.

### Associations between grey matter volume and PCA-derived factor scores

The neuroanatomical correlates of neuropsychological performance in FTD were explored using voxel-based correlational methodology (VBCM).^87^ A linear regression model was fitted to explore the association between grey matter intensity and factor scores on each neuropsychological component, with age, ICV and scanner site included as covariates. Significant clusters were extracted using a cluster-level threshold of *Q* < 0.05, based on an initial voxel-level threshold of *P* < 0.001. To explore the contributions of not only ATL magnitude, but also ATL asymmetry and OFC magnitude/asymmetry, forced-entry multiple linear regression models were fitted to predict scores on each neuropsychological task, with the four ROIs as predictors.

## Results

### Neuroimaging comparisons

#### Grey matter volume differences between groups

The VBM results align closely with the expected patterns for each clinical group (Fig. 1 and Supplementary Table 1). Direct comparisons between FTD subgroups revealed reduced grey matter in the bilateral ATLs in SD (Supplementary Fig. 2) and no significant clusters for the reverse contrast. As expected, the resected TLE patients provide a neuroanatomical model of (i) purely unilateral and complete resection and (ii) no detected frontal changes – which is a powerful comparison to the concurrent frontal, temporal and insular atrophy of the FTD patients. These patterns were underlined by the ROI analyses (see next).

**Figure 1.**
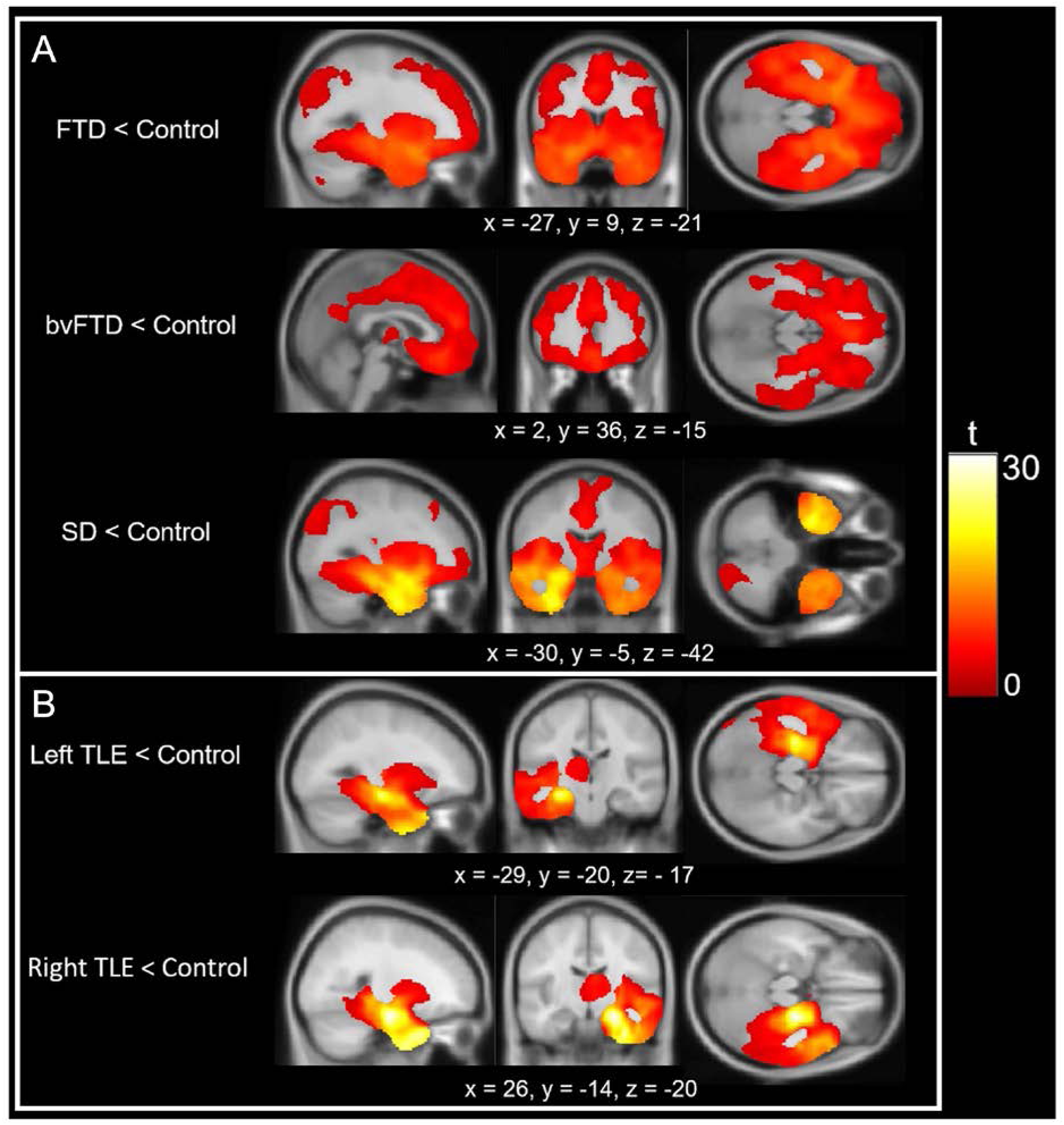
Voxel-based morphometry results. Each row displays clusters of reduced grey matter volume compared to age-matched controls for **(A)** FTD and **(B)** TLE. Groups were compared using independent t-tests, with age, intracranial volume and scanner site included as covariates. Images are thresholded using a cluster-level threshold of *Q* < 0.05 (after an initial voxel-level threshold of *P* < 0.001). Significant clusters are overlaid on the MNI avg152 T1 template. Co-ordinates are reported in Montreal Neurological Institute space.

#### Magnitude and asymmetry in core frontotemporal regions

Grey matter indices for each participant are displayed in Fig. 2 and reported in Table 2. There was a significant group difference in ATL magnitude (*F*(3, 46) = 14.60, *P* < 0.0001), with post-hoc tests revealing that the SD group had lower magnitude than bvFTD (*P* < 0.0001) and left TLE (*P* < 0.0001), but not right TLE (*P* = 0.37), whereas the right TLE group had lower magnitude compared to bvFTD (*P* = 0.03). Groups also differed in ATL asymmetry (*F*(3, 46)

**Figure 2.**
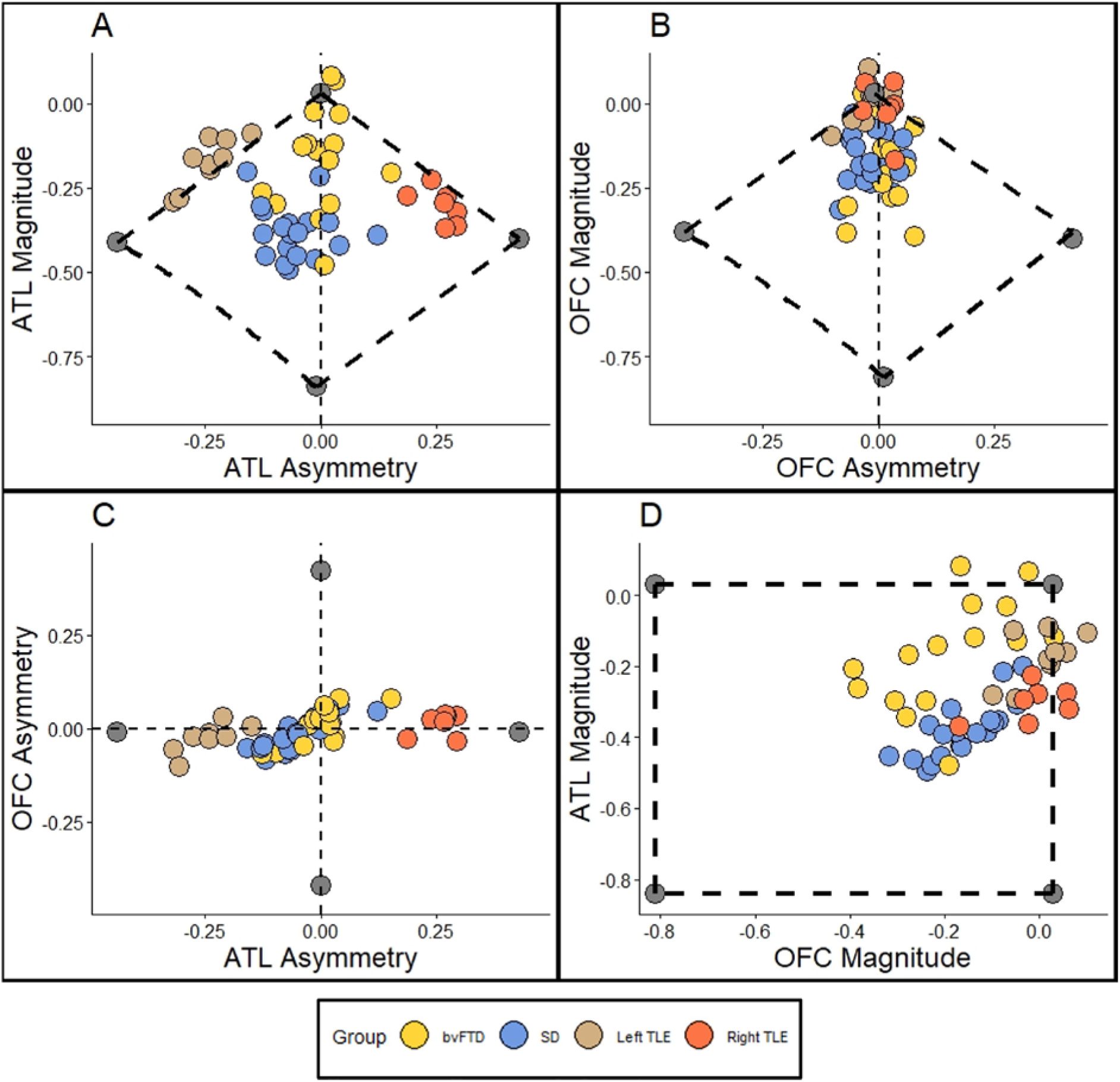
Scatter plots displaying ATL and OFC indices for each patient. Lower magnitude values indicate greater volume loss and negative asymmetry values indicate left > right volume loss. **(A)** ATL magnitude vs. ATL asymmetry**. (B)** OFC magnitude vs. OFC asymmetry. **(C)** OFC asymmetry vs. ATL asymmetry. **(D)** ATL magnitude vs. OFC magnitude. The grey points represent the extremity boundaries.

**Table 2.** Magnitude and asymmetry grey matter indices for each group.

|  | **bvFTD** | **SD** | **Left TLE** | **Right TLE** | **Group difference** | **Post-hoc** |
| --- | --- | --- | --- | --- | --- | --- |
| N | 15 | 19 | 9 | 7 | - | - |
| ATL Magnitude | -0.17 (0.16) | -0.38 (0.08) | -0.18 (0.07) | -0.30 (0.05) | **F(3,46)=14.60, P < 0.0001** | SD, R < bvFTD; SD < L |
| ATL Asymmetry (absolute value) | 0.04 (0.05) | 0.07 (0.05) | 0.24 (0.05) | 0.26 (0.04) | **F(3,46)=64.78, P < 0.0001** | bvFTD, SD < L, R |
| OFC Magnitude | -0.19 (0.13) | -0.16 (0.08) | 0.007 (0.06) | -0.02 (0.08) | **F(3,46) = 12.19, P < 0.0001** | bvFTD, SD < L, R |
| OFC Asymmetry (absolute value) | 0.04 (0.03) | 0.04 (0.02) | 0.03 (0.03) | 0.03 (0.006) | F(3,46)=0.46, P = 0.72 | - |
| ATL Magnitude vs. ATL Asymmetry | $r = 0.21$ | $r = -0.19$ | **$r = 0.85^{**}$** | $r = -0.57$ | - | - |
| ATL Magnitude vs. OFC Magnitude | **$r = 0.54^*$** | **$r = 0.78^{****}$** | $r = 0.58$ | $r = 0.48$ | - | - |
| ATL Magnitude vs. OFC Asymmetry | $r = -0.03$ | $r = -0.09$ | **$r = 0.68^*$** | $r = 0.21$ | - | - |
| ATL Asymmetry vs. OFC Magnitude | $r = 0.10$ | $r = -0.12$ | $r = 0.57$ | $r = -0.25$ | - | - |
| ATL Asymmetry vs. OFC Asymmetry | **$r = 0.74^{**}$** | **$r = 0.83^{****}$** | **$r = 0.76^*$** | $r = 0.35$ | - | - |
| OFC Asymmetry vs. OFC Magnitude | $r = -0.10$ | $r = 0.009$ | **$r = 0.67^*$** | $r = -0.34$ | - | - |

Top four rows display mean and standard deviations for each group. Bottom four rows display Pearson's correlation coefficients between each index. Significant p-values are highlighted in bold.
\* $P < 0.05$ ; \*\* $P < 0.01$ ; \*\*\* $P < 0.001$ ; \*\*\*\* $P < 0.0001$
ATL = anterior temporal lobe, bvFTD = behavioural-variant frontotemporal dementia, C = control, L = left TLE, OFC = orbitofrontal cortex, R = right TLE, SD = semantic dementia

= 64.78, *P* < 0.0001). Both TLE groups had significantly greater absolute asymmetry values than both bvFTD and SD (*P* < 0.0001). There was a significant group effect on OFC magnitude (*F*(3, 46) = 12.19, *P* < 0.0001). As expected, both FTD subgroups had significantly lower magnitude than TLE (all *P* < 0.01), with no significant differences between bvFTD and SD (*P* = 0.83) or between left and right TLE (*P* = 0.96). There was no main effect of group on OFC asymmetry (F(3, 46) = 0.46, *P* = 0.72). ATL and OFC magnitude were positively correlated in both FTD subgroups (bvFTD; *r* = 0.54, *P* = 0.04, SD; *r* = 0.78, *P* < 0.0001), but not in TLE (left TLE; *r* = 0.58, *P* = 0.10, right TLE; *r* = 0.48, *P* = 0.27). Asymmetry indices also were strongly positively correlated in bvFTD (*r* = 0.74, *P* = 0.002), SD (*r* = 0.83, *P* < 0.0001) and left TLE (*r* = 0.76, *P* = 0.03), although not in right TLE (*r* = 0.35, *P* = 0.44).

### Neuropsychological comparisons

#### General semantic memory and background neuropsychology

Table 3 displays scores for each group on each test. Both FTD subgroups were impaired on every semantic task and each ACE-R subscale (*P* < 0.05). Relative to bvFTD, the SD group performed more poorly on the Cambridge (*P* = 0.0006) and Boston (*P* = 0.001) Naming tests, and the ACE-R Language subscale (*P* = 0.008) and the bvFTD group had lower scores than SD on the Raven’s (*P* = 0.006). The left TLE group were impaired on the Boston Naming test (*P* = 0.02), synonym judgement task (*P* = 0.002), as well as the Memory (*P* = 0.03), Fluency (*P* = 0.04) and Language (*P* = 0.04) ACE-R subscales. There were no significant differences between left and right TLE on any tasks.

**Table 3.** Mean scores on each task.

|  | Control | bvFTD | SD | Left<br>TLE | Right<br>TLE | Group difference | Post-hoc |
| --- | --- | --- | --- | --- | --- | --- | --- |
| N | 19 | 26 | 22 | 11 | 7 | - | - |
| ACE-R Total (100) | 96.8 (2.3) | 60.2 (22.0) | 45.4 (23.5) | 80.5 (9.8) | 87.7 (6.1) | **H(4) = 58.9\*\*** | L, bvFTD, SD < C<br>SD < L<br>bvFTD, SD < R |
| MMSE (30) | 29.8 (0.4) | 21.3 (6.9) | 19.1 (8.9) | 27.2 (1.5) | 28.9 (1.1) | **H(4) = 50.8\*\*** | L, bvFTD, SD < C<br>bvFTD, SD < R |
| ACE-R Attention (18) | 17.9 (0.2) | 13.4 (4.7) | 12.5 (5.8) | 17.4 (0.9) | 17.9 (0.4) | **H(4) = 41.0\*\*** | bvFTD, SD < C, L, R |
| ACE-R Memory (26) | 24.5 (2.0) | 13.2 (7.9) | 8.1 (6.5) | 16.6 (5.5) | 19.9 (4.4) | **H(4) = 45.2\*\*** | L, bvFTD, SD < C<br>SD < R |
| ACE-R Fluency (14) | 13.2 (1.2) | 4.0 (3.3) | 4.5 (3.4) | 9.1 (1.9) | 11.0 (1.9) | **H(4) = 59.0\*\*** | L, bvFTD, SD < C<br>bvFTD, SD < L<br>bvFTD, SD < R |
| ACE-R Language (26) | 25.7 (0.5) | 18.2 (7.1) | 8.8 (5.4) | 21.9 (3.8) | 23.4 (1.7) | **H(4) = 57.5\*\*** | L, bvFTD, SD < C<br>SD < L<br>SD < R<br>SD < bvFTD |
| ACE-R Visuospatial (16) | 15.6 (0.8) | 11.3 (3.8) | 11.5 (4.9) | 15.5 (0.8) | 15.6 (0.5) | **H(4) = 31.8\*\*** | bvFTD, SD < C, L, R |
| Cambridge Naming (32) | 31.9 (0.2) | 27.3 (7.8) | 13.0 (9.7) | 31.2 (1.4) | 31.9 (0.4) | **H(4) = 57.5\*\*** | bvFTD, SD < C<br>SD < L<br>SD < R<br>SD < bvFTD |
| Boston Naming (30) | 29.7 (0.5) | 21.5 (8.5) | 6.8 (5.5) | 26.1 (2.4) | 27.9 (2.3) | **H(4) = 60.8\*\*** | L, bvFTD, SD < C<br>SD < L, R, bvFTD |
| Camel and cactus test (32) | 30.7 (1.1) | 21.8 (7.4) | 15.7 (5.1) | 28.8 (1.8) | 29.0 (1.6) | **H(4) = 47.2\*\*** | bvFTD, SD < C<br>SD < L<br>SD < R |
| Synonym judgement (48) | 47.8 (0.4) | 39.0 (7.9) | 35.9 (7.5) | 42.9 (1.8) | 44.9 (2.9) | **H(4) = 47.5\*\*** | L, bvFTD, SD < C<br>SD < R |
| Raven's (12) | 10.5 (1.5) | 5.0 (2.7) | 8.3 (3.4) | 10.2 (1.4) | 10.3 (1.9) | **H(4) = 35.8\*\*** | bvFTD < C, L, R, SD |
| Brixton (10) | 6.4 (2.0) | 2.9 (2.0) | 4.8 (2.8) | 6.6 (2.2) | 5.7 (2.0) | **H(4) = 25.5\*\*** | bvFTD < C, L |
| Face-name matching (44) | 38.9 (3.4) | 28.6 (11.1) | 16.4 (7.9) | 36.8 (4.2) | 34.3 (10.1) | **H(4) = 34.8\*\*** | bvFTD, SD < C<br>SD < L, R, bvFTD |
| Face-profession matching (44) | 40.3 (3.7) | 27.7 (11.4) | 20.0 (10.0) | 39.6 (3.3) | 37.3 (6.9) | **H(4) = 35.2\*\*** | bvFTD, SD < C, L<br>SD < R |
| Landmark-name matching (42) | 38.5 (1.8) | 24.3 (9.0) | 16.1 (6.3) | 27.2 (3.6) | 30.9 (5.9) | **W(4, 24) = 77.2\*\*** | bvFTD, SD, L < C<br>SD < L, R, bvFTD |
| Famous face matching (22) | 21.2 (0.8) | 18.6 (2.8) | 18.0 (2.1) | 20.6 (2.0) | 18.9 (2.4) | **H(4) = 26.7\*\*** | bvFTD, SD < C<br>SD < L |
| Unfamiliar face matching (22) | 20.3 (1.4) | 17.1 (3.1) | 18.1 (2.8) | 19.1 (1.2) | 16.7 (2.3) | **H(4) = 19.1\*** | bvFTD, SD, R < C |
| Social abstract synonym<br>judgement (36) | 33.9 (1.3) | 26.4 (5.3) | 25.1 (5.8) | 31.1 (1.3) | 32.3 (2.1) | **H(4) = 42.8\*\*** | bvFTD, SD < C<br>SD < R |
| Non-social abstract synonym<br>judgement (36) | 35.6 (0.6) | 28.1 (6.1) | 25.5 (6.7) | 33.1 (3.0) | 34.9 (1.2) | **H(4) = 42.2\*\*** | bvFTD, SD < C<br>SD < R |
| Social roles word-picture<br>matching (35) | 34.4 (0.8) | 29.8 (5.8) | 27.1 (6.0) | 32.5 (1.1) | 32.9 (1.3) | **H(4) = 40.2\*\*** | bvFTD, SD < C |
| Non-social word-picture<br>matching (36) | 35.9 (0.2) | 33.5 (5.7) | 29.3 (6.9) | 35.8 (0.4) | 36.0 (0.0) | **H(4) = 35.6\*\*** | SD < C, L, R, bvFTD |
| Basic emotion matching (19) | 16.3 (1.5) | 11.8 (3.2) | 11.0 (3.5) | 15.3 (1.7) | 14.7 (1.8) | **H(4) = 36.0\*\*** | bvFTD, SD < C<br>SD < L |
| Complex emotion matching (23) | 18.5 (1.9) | 12.9 (4.9) | 12.2 (5.0) | 16.9 (2.1) | 17.4 (3.9) | **W(4, 24.4) = 9.5\*\*** | bvFTD, SD < C, L |
| Social Norms Questionnaire<br>(22) | 20.0 (1.2) | 15.4 (3.9) | 15.5 (2.7) | 19.4 (1.1) | 19.9 (0.7) | **H(4) = 32.5\*\*** | bvFTD, SD < C, L, R |
| TASIT-Sarcasm (20) | 18.7 (1.9) | 9.9 (5.5) | 8.1 (5.1) | 15.3 (2.7) | 15.0 (2.7) | **H(4) = 38.0\*\*** | bvFTD, SD < C |

Means and standard deviations for each group reported. \* $P < 0.001$ ; \*\* $P < 0.0001$ . ACE-R = Addenbrooke's Cognitive Examination-Revised, bvFTD = behavioural-variant frontotemporal dementia, C = control, L = left TLE, MMSE = Mini Mental State Examination, R = right TLE, SD = semantic dementia, TASIT = The Awareness of Social Inference Test, TLE = temporal lobe epilepsy

#### Social-semantic battery

FTD groups were impaired across all tasks in the social-semantic battery (*P* < 0.05), the only exception being bvFTD on the non-social word-picture matching task (*P* = 0.22). Direct comparisons between FTD subtypes found that the SD group performed more poorly on non-social word-picture matching (*P* = 0.004), face-name matching (*P* = 0.03) and landmark-name matching (*P* = 0.01). The left TLE group were impaired on the landmark-name matching (*P* < 0.0001), whereas the right TLE group were impaired on unfamiliar perceptual face matching (*P* = 0.005). As with the general semantic tasks, there were no significant differences between left and right TLE.

### Single-case explorations

Twenty-two participants (bvFTD = 10, SD = 10, left TLE = 2) fulfilled criteria for a strong dissociation. In every case, this was due to poorer performance on the abstract non-social synonym task compared to the social task. Only one participant met criteria for a classical dissociation; an SD patient who was selectively impaired on non-social concepts (*P* = 0.0002). In summary, there was no evidence from the single case explorations for any neuropsychological dissociations in either FTD or ATL-resected TLE. Scores for the FTD and TLE participants on the two tasks are displayed in Supplementary Fig. 3.

### Extracting neuropsychological components

The percentage of participants too impaired to complete each task and hence scored at chance-level is reported in Supplementary Table 2. The KMO statistic was 0.87, indicating meritorious sampling adequacy,^88^ and Bartlett’s test for sphericity was significant (*P* < 0.0001), indicating presence of at least some common factors in the covariance matrix. Visual inspection of the scree plot indicated three principal components (Supplementary Fig. 4) which explained 78.5% of the total variance.

Task and factor loadings are displayed in Fig. 3. The first principal component (PC) had an eigenvalue of 14.5 and explained 60.2% of the total variance. The tasks loading positively onto this component were ACE-R Attention, ACE-R Visuospatial, ACE-R Fluency, CCT, synonym judgement, Raven’s, social word-picture matching, non-social word-picture matching, famous and unfamiliar perceptual face matching, emotion matching, and the abstract social and non-social synonym judgement tasks. There is no specific cognitive process shared by all tasks, but rather this component reflects *FTD severity* – in keeping with sampling FTD specifically (rather than many kinds of dementia or aetiologies) and testing them on a collection of tasks known to be affected in this group. In keeping with this interpretation, scores on this factor were strongly correlated with total atrophy across the patients while the other factors were not (Supplementary Fig. 5). There were no statistically reliable differences in mean factor scores between bvFTD and SD on this component (*t* = 0.44, *P* = 0.66).

**Figure 3.**
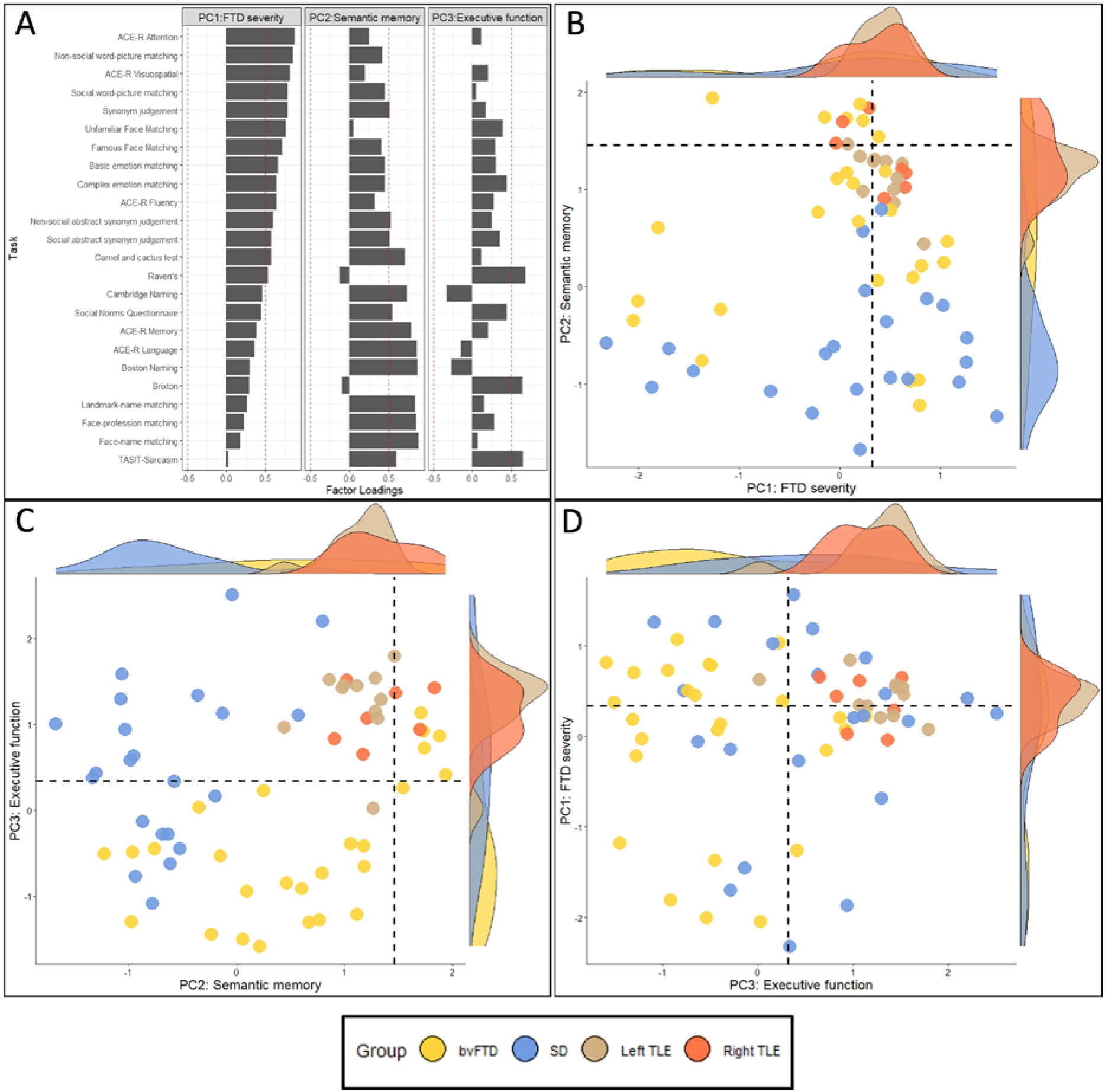
PCA factor loadings and factor scores. (A) Factor loadings for each task. Red dashed lines indicate factor loading cut-offs (>|0.5|). **(B)** PC1 (FTD severity) plotted against PC2 (semantic memory). **(C)** PC2 (semantic memory) plotted against PC3 (executive function). **(D)** PC3 (executive function) plotted against PC1 (FTD severity). The black dashed lines indicate the factor score of a hypothetical participant scoring 1.96 standard deviations below the control average on each task.

The second PC had an eigenvalue of 2.99 and explained 12.5% of the remaining variance. Tasks loading positively were the ACE-R Memory, ACE-R Language, Cambridge Naming, Boston Naming, CCT, synonym judgement, face-name matching, landmark-name matching, SNQ and abstract social and non-social synonym judgement tasks. This component was labelled *semantic memory* as it primarily included semantic tasks. The SD group had significantly lower factor scores (i.e. poorer performance) on this component compared to bvFTD (*t* = 5.38, *P* < 0.0001). Crucially, both social *and* non-social semantic tasks co-loaded onto this component, and thus we use ‘semantic memory’ to refer to both ‘social- and non-social-semantic memory’. Indeed, when we extracted separate ‘social’ and ‘non-social’ factors (by entering the two sets of assessment results into two separate one-factor PCAs), scores on these two factors were highly correlated (*r* = 0.85, *P* < 0.0001), which strongly suggests the generalised degradation of a unitary conceptual system affecting both social and non-social concepts in FTD.

The third PC had an eigenvalue of 1.41 and explained 5.9% of the remaining variance. Tasks loading positively were the two executive function tasks and the TASIT-sarcasm. Consequently, this PC was labelled *executive function*. The bvFTD group had significantly lower factor scores on this component than SD (*t* = 3.97, *P* = 0.0002).

#### Projection of TLE participants into the FTD-defined PCA space

The TLE patients’ neuropsychological scores were projected into the FTD-defined PCA space using the regression method (Fig. 3). We then used ANOVAs to assess whether the TLE groups differed from bvFTD and SD in their average scores on each factor. There was no significant effect of group on *FTD severity* factor scores (*F*(3, 62) = 1.05, *P* = 0.38), but there was a large group effect on *semantic memory* factor scores (*F*(3, 62) = 24.14, *P* < 0.0001) with SD having lower scores than both TLE groups (*P* < 0.0001), as well as a large effect on *executive function* factor scores (*F*(3, 62) = 16.74, *P* < 0.0001) where the bvFTD scores were lower than both TLE groups (*P* < 0.0001). Most of the left (90.9%) and right (57.1%) TLE participants had a *semantic memory* factor score below the control-derived cut-off (defined as the factor score of a hypothetical participant scoring 1.96 SDs below the control average on all tasks), but no TLE participant had a factor score below the cut-off for *executive function*. There were no differences between left and right TLE on *FTD severity* (*P* = 0.99), *semantic memory* (*P* = 0.93) or *executive function* (*P* = 0.99). Taken together, these findings suggest that unilateral ATL resection yields a mild generalised semantic impairment in the context of preserved executive function, with no clear left vs. right differences.

#### Association between grey matter volume and neuropsychological performance

*FTD severity* factor scores were associated with grey matter volume in the precentral gyrus, frontal/orbital gyri, cingulate cortex, insula and supplementary motor area (Fig. 4A). Reinforcing the interpretation of PC1 as representing *FTD severity*, changes of grey matter in a very similar set of regions were found to correlate with levels of global atrophy (Fig. 4B). Indeed, (i) total grey matter volume and *FTD severity* scores were found to be strongly correlated (*r* = 0.46; *P* = 0.006), and (ii) when total grey matter volume was entered as a covariate into the FTD severity VBCM analysis then no regions remained. *Semantic memory* factor scores were associated with grey matter volume in the bilateral ATLs, maximal at the temporal poles and ventral ATL regions (Fig. 4C). These semantic-to-atrophy correlations were unchanged when total atrophy was entered as a covariate (and the semantic PCA scores were not significantly correlated with global atrophy: *r* = 0.31, *P* = 0.07). No significant clusters emerged for *executive function*.

**Figure 4.**
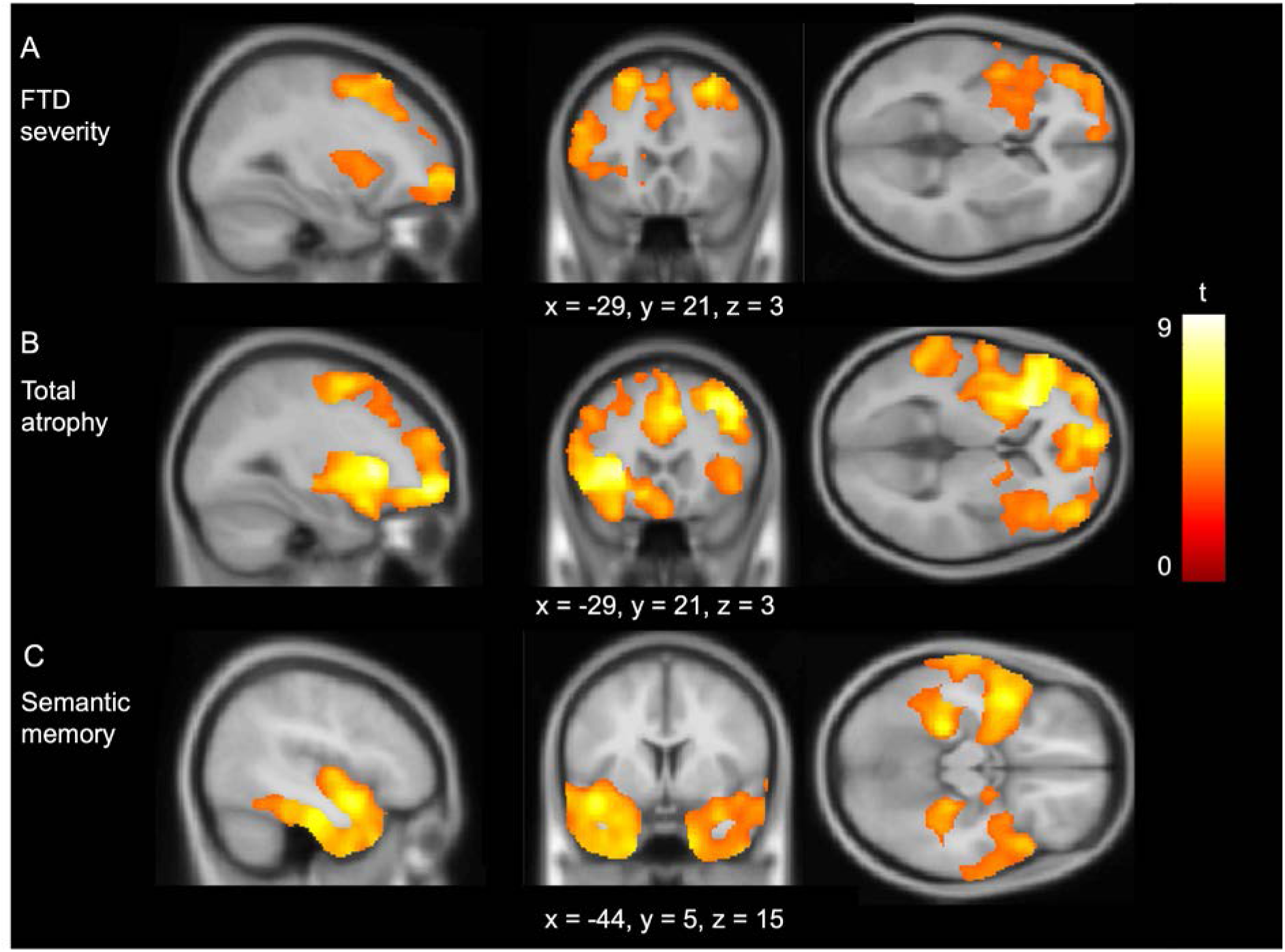
Regions of grey matter volume associated with factor scores. Regions of grey matter positively correlated with **(A)** FTD severity factor scores, **(B)** Total grey matter volume and **(C)** Semantic memory factor scores. Multiple linear regression models were fitted with each factor as the main effect, with age, intracranial volume and scanner site included as covariates. Images are thresholded using a cluster-level threshold of *Q* < 0.05 (above an initial voxel-level threshold of *P* < 0.001). Significant clusters are overlaid on the MNI avg152 T1 template. Co-ordinates are reported in Montreal Neurological Institute space.

To explore the importance of the bilateral and/or asymmetric nature of the atrophy, linear multiple regression models were fitted with the ATL and OFC indices as predictors. The model was significant for *semantic memory* factor scores (*F*(4, 29) = 18.30, *P* < 0.0001) with the magnitude of ATL atrophy the only significant individual predictor (*t* = 7.82, *P* < 0.0001). However, ATL asymmetry was not significant (*t* = 0.29, *P* = 0.78). The linear multiple regression model was significant for ACE-R Memory (*F*(4, 29) = 7.05, *P* = 0.0004), ACE-R Language (*F*(4, 29) = 12.97, *P* < 0.0001), Cambridge Naming (*F*(4, 29) = 8.56, *P* = 0.0001), Boston Naming (*F*(4, 29) = 15.06, *P* < 0.0001), face-name matching (*F*(4, 25) = 8.22, *P* = 0.0002), face-profession matching (*F*(4, 24) = 10.69, *P* < 0.0001) and landmark-name matching (*F*(4, 25) = 5.09, *P* = 0.004). ATL magnitude was the only significant predictor in every case, except for the landmark-name matching (also predicted by OFC magnitude; *t* = -2.65, *P* = 0.01) and the two naming tasks which were also predicted by ATL asymmetry (Cambridge Naming; *t* = 2.16, *P* = 0.04, Boston Naming; *t* = 2.17, *P* = 0.04). Full details of each regression model are reported in Table 4.

**Table 4.**
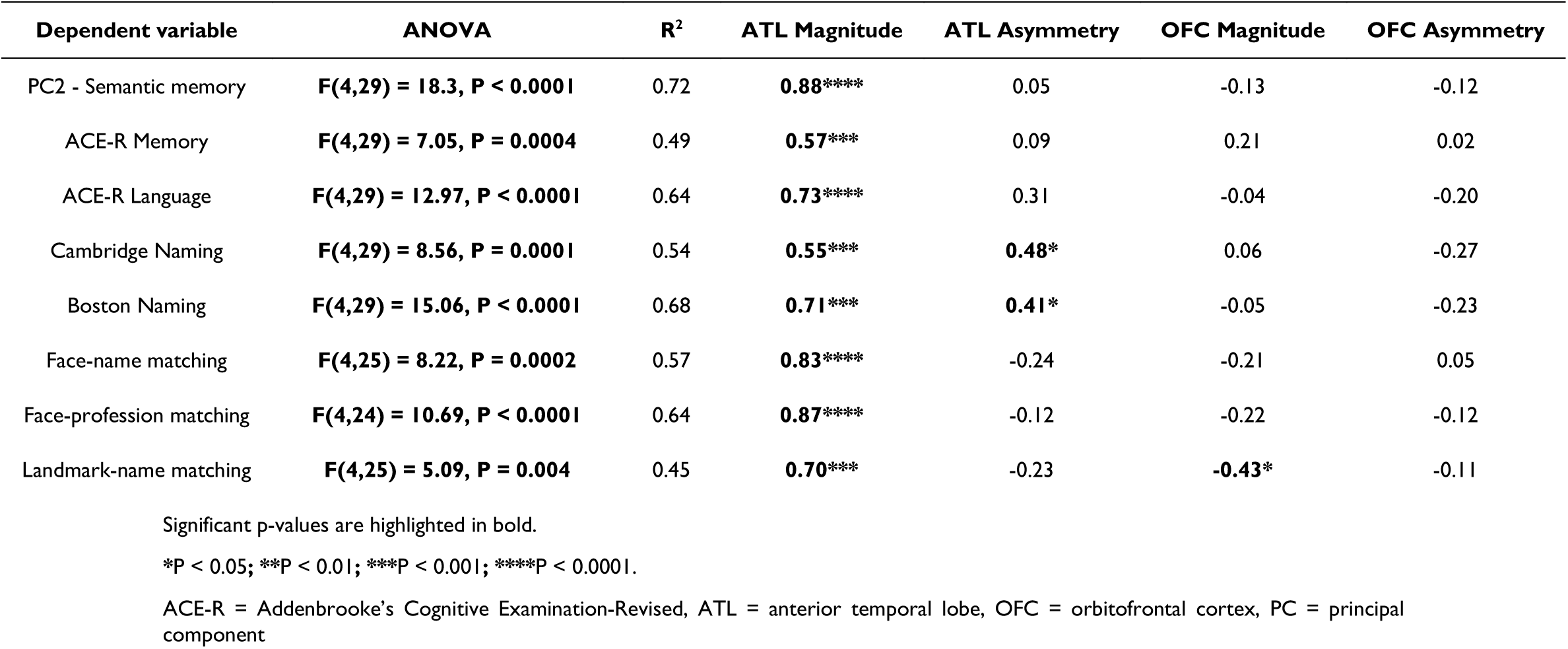
Model summaries and standardised beta-values for each regression model.

| Dependent variable | ANOVA | R<sup>2</sup> | ATL Magnitude | ATL Asymmetry | OFC Magnitude | OFC Asymmetry |
| --- | --- | --- | --- | --- | --- | --- |
| PC2 - Semantic memory | **F(4,29) = 18.3, P < 0.0001** | 0.72 | **0.88\*\*\*\*** | 0.05 | -0.13 | -0.12 |
| ACE-R Memory | **F(4,29) = 7.05, P = 0.0004** | 0.49 | **0.57\*\*\*** | 0.09 | 0.21 | 0.02 |
| ACE-R Language | **F(4,29) = 12.97, P < 0.0001** | 0.64 | **0.73\*\*\*\*** | 0.31 | -0.04 | -0.20 |
| Cambridge Naming | **F(4,29) = 8.56, P = 0.0001** | 0.54 | **0.55\*\*\*** | **0.48\*** | 0.06 | -0.27 |
| Boston Naming | **F(4,29) = 15.06, P < 0.0001** | 0.68 | **0.71\*\*\*** | **0.41\*** | -0.05 | -0.23 |
| Face-name matching | **F(4,25) = 8.22, P = 0.0002** | 0.57 | **0.83\*\*\*\*** | -0.24 | -0.21 | 0.05 |
| Face-profession matching | **F(4,24) = 10.69, P < 0.0001** | 0.64 | **0.87\*\*\*\*** | -0.12 | -0.22 | -0.12 |
| Landmark-name matching | **F(4,25) = 5.09, P = 0.004** | 0.45 | **0.70\*\*\*** | -0.23 | **-0.43\*** | -0.11 |

Significant p-values are highlighted in bold.
\*P < 0.05; \*\*P < 0.01; \*\*\*P < 0.001; \*\*\*\*P < 0.0001.
ACE-R = Addenbrooke's Cognitive Examination-Revised, ATL = anterior temporal lobe, OFC = orbitofrontal cortex, PC = principal component

## Discussion

This study considered how *social-semantic knowledge* is (a) impaired in FTD relative to general semantic memory and (b) differentially supported by the left *vs.* right ATLs. We conducted a comprehensive and systematic investigation of social concepts using a battery comprising diverse types of social concept and non-social-semantic tasks. The results suggest that semantic knowledge in both social and non-social domains are equally affected by ATL damage, with little difference between left- vs. right-predominant abnormality in either domain. People who had undergone unilateral ATL resection provided convergent data supporting this conclusion. In the following sections, we discuss the key findings and clinical implications.

### Social and non-social concepts are underpinned by the bilateral anterior temporal lobes

A selective degradation of conceptual knowledge is the defining feature of SD.^2,3,5,6^ Research over recent decades has revealed that this degradation occurs for all types of concepts, in their verbal and non-verbal modalities, following bilateral ATL atrophy.^4,18,21,22^ In this study, we have demonstrated that the conceptual degradation extends to a very wide range of social concepts. Although milder than in SD, a parallel decline in social- and non-social-semantic knowledge was also found in bvFTD, highlighting the phenotypic overlap between the syndromes and mirroring the neuroanatomical overlap including ATL atrophy.^8,9,89,90^ Indeed, a very clear picture emerges by adopting a transdiagnostic approach: the PCA conducted across SD and bvFTD patients indicated that both social- and non-social-semantic deficits were highly correlated and heavily co-loaded onto the same *semantic memory* component. Factor scores were associated with grey matter volume only in the bilateral ATLs when the entire FTD group was analysed together. This is true not only in the SD subset of cases (i.e., the classical ATL-semantically impaired patient population) but also in the remaining FTD patients (i.e., when a patient with more frontally-centred atrophy presents with a semantic impairment, this is due to concurrent ATL atrophy, rather than representing a distinct new subtype of bvFTD). In addition, the ROI regression analyses showed that semantic scores were associated with total <u>bilateral</u> ATL volume, but *not* ATL asymmetry. Indeed, ATL asymmetry was not associated with performance on any individual social-semantic comprehension task. These findings demonstrate that social- and non-social-semantic knowledge is supported by the ATLs bilaterally. There was no evidence (i) that social-semantic knowledge is neuroanatomically distinct from general conceptual knowledge or (ii) that R>L ATL atrophy causes increased social-semantic impairments relative to L>R atrophy. It is important to note that the analyses of this large dataset were able to detect asymmetrically supported functions where they did occur: as found in previous studies of SD and many other patient groups,^1,53,91,92^ plus in healthy participants after rTMS, naming and speech production are substantially more affected by damage/stimulation to the left than right ATL.^93^ Past neuroanatomically-constrained computational models have shown that this follows as a corollary of a bilaterally-supported ATL semantic system driving left-lateralised speech production.^92,94^

Unilateral ATL resection yielded mild impairments across social- and non-social-semantic tasks, and on the PCA semantic memory factor score. These findings replicate previous studies, where unilateral ATL damage is associated with a mild semantic impairment when sensitive assessments are used.^49,53–55^ The chasm between the subtle unilateral and severe bilateral effects on semantic processing cannot be explained solely by the degree of total ATL damage, as many of the TLE participants had a magnitude of ATL grey matter loss similar to that in some cases of SD (see Fig. 2). In other words, although the level of semantic impairment in these patients is clearly governed by the overall amount of ATL damage, the distribution of damage across the left and right ATLs is also crucial. Bilateral-implementation may configure the semantic system to be resilient to unilateral damage, a hypothesis that has been formally captured and explored computationally.^94^ This investigation found that when the semantic representation “ATL” hub was divided into partially interconnected “demi-hubs” (mimicking the left and right ATLs), simulated unilateral damage caused a much milder impairment than bilateral damage, even when levels of total damage were kept the same. Following unilateral damage, the intact contralateral demi-hub was able to function with higher accuracy, albeit slower than before.^94^ Formal analyses of this model showed there were two causes of this bilateral versus unilateral discrepancy. First, there is some redundancy of semantic representation across the “demi-hubs”. Second, damage generates noise which propagates to connected units: after unilateral damage this noise propagation is constrained to one demi-hub, whereas after bilateral damage this noise percolates throughout the system.

Secondary to the mild generalised semantic impairment, graded neuropsychological differences can emerge from left vs. right ATL unilateral damage. Consistent with the results from SD and associated computational models (see above), increased anomia is found after left ATL resection.^53,91^ Despite left versus right differences for naming and perceptual face matching, we found no evidence of any differences in social (or non-social) semantics in the surgical cases – again mirroring the findings from FTD. Moreover, in contrast to the right ATL hypothesis for social processing, the TLE participants (right and left) show no behavioural changes, even when formally assessed using the same neuropsychiatric tools as those used in FTD.^53^

In contrast to FTD, where the atrophy occurs in the context of a previously intact and typically organized semantic system, the chronic epilepsy in TLE raises the possibility of pre-surgical functional reorganisation away from seizure centres. This idea is potentially consistent with findings of functional and structural connectivity changes to language-networks in TLE.^95–97^ Under this commonly-rehearsed hypothesis, the minimal impact of resection on semantic memory may occur because the ATL is no longer supporting this function. This is an important potential confound to consider when interpreting differences between ATL-resected TLE and FTD. However, a moment of further reflection on this idea is merited for multiple reasons and new data. Functional re-organization in TLE is associated with young-onset epilepsy (presumably due to greater capacity for plasticity in the developing brain)^95,98,99^ whereas the TLE participants in the current study were adult-onset, suggesting an increased likelihood of typical ATL organization. Furthermore, there is evidence that any such reorganisation is minimal, at least for semantic representation. First, as described above, very mild generalised semantic impairments are found in unilateral ATL-resected cases^49,53–55^ and the degree of semantic impairment is associated with the amount of resected tissue.^55^ Second, the increased anomia caused by left ATL resection mimics the relatively more severe anomia in L>R SD,^92^ which implies that semantic memory is organised similarly in pre-surgical TLE as in SD. Third, direct cortical grid electrode studies of pre-surgical TLE patients detect semantic-related neural activity in the left and right ventrolateral ATLs and cortical stimulation generates a transient semantic impairment in exactly the same semantic “hot-spot” as that observed in healthy participant fMRI studies.^20,100,101^ Finally, task-based fMRI in resected TLE patients shows that, rather than shifts of semantic function to new locations, the patients’ semantic system upregulates activation in the same (remaining) regions as those observed in healthy participants^102^; this pattern is closely mirrored in healthy participants after rTMS to the ATL.^103,104^

The PCA extracted three components. We labelled the first *FTD severity,* as it included a range of tasks with no specific underlying cognitive process. In keeping with this interpretation, bvFTD and SD patients had similar factor scores on this component, and factor scores were correlated with overall grey matter volume. The two remaining components had more precise neuropsychological labels and corresponded to *semantic memory* and *executive function* respectively. Multiple tasks loaded across more than one component; each component included a mixture of social- and non-social measures, as well as tasks not generally characterized as primarily ‘semantic’ or ‘executive’. This highlights that no one task is ‘pure’, but instead draws on multiple cognitive elements which adds complexity to the interpretation of PCA-derived factors. The PCA distills the core underlying cognitive dimensions in the data, with task loadings indicating how each task decomposes across these dimensions. Accordingly, some tasks loaded singly onto certain factors (e.g., naming on *semantic memory*), whereas others loaded across multiple components (e.g., TASIT-sarcasm on *semantic memory* and *executive function,* indicating that performance on this task requires both conceptual and executive processing).

### Clinical implications

#### Social-semantic knowledge and ‘right’ semantic dementia

FTD patients with R>L ATL atrophy often present to clinic with behavioural changes.^42–44^ This clinical observation has driven the hypothesis that the right ATL has a specialised role in social processing^14,16^ and proposals that R>L SD is a distinct clinical syndrome.^14,42,45^ Our results challenge this view. From both FTD and resected TLE, we found no evidence of right-lateralised specialisation for social concepts, but rather equal contributions from left and right ATL to all types of semantic knowledge. As noted above, this finding aligns with parallel fMRI and rTMS ATL explorations in healthy participants.^32,33,58,59^ What, then, is the cause of the commonly observed social problems in patients with R>L SD? R>L cases typically present to clinic later than L>R, and even though they must exist, there is a paucity of early R>L SD patients in the literature, either as single cases or as part of group studies, including the current investigation (for a review, see^1^). Group studies have found that R>L SD patients typically have more overall temporal lobe atrophy than L>R^1,42^ and increased prefrontal atrophy.^51^ There are at least three (non-mutually exclusive) alternative explanations for the increased behavioural change in R>L SD. First, R>L SD cases have greater overall ATL volume loss, bilaterally, which would cause a relatively greater degradation of semantic memory (for both social and non-social concepts) which is important for supporting appropriate social behaviour.^13^ Second, the increased behavioural changes result from increased prefrontal damage in areas important for controlled social behaviour, such as the OFC.^52^ Third, we demonstrated that ATL and OFC asymmetry are correlated in FTD, raising the possibility that R>L OFC asymmetry may also contribute to the increased behaviour change. Indeed, theories of behavioural change in FTD have highlighted the importance of right prefrontal regions, in particular, in social functioning.^105^

Data-driven analyses revealed that L > R and R > L cases had highly overlapping neuropsychological profiles, and single-case explorations found no evidence for selective social deficits in R > L patients. Instead, L >R and R >L patients had a dual social and non-social conceptual degradation following bilateral ATL atrophy. Although there was some variations in asymmetry, the ATL atrophy in SD was highly bilateral compared to the unilateral ATL-resected participants. Based on these findings, we conceptualize SD as a *unitary-yet-graded* disorder, where asymmetric L > R (svPPA) and R > L (‘right’ SD) represent points on a spectrum (that become increasingly similar over time as atrophy rapidly spreads into the contralateral ATL).^1,106^ In keeping with this proposal, L > R and R >L cases are both associated with TDP-43 type C pathology, suggesting they reflect presentations of the same disease.^107^

#### A transdiagnostic approach to frontotemporal dementia

Although there were broad group level differences in keeping with the paradigmatic phenotypes of each FTD syndrome (i.e., poorer semantic memory in SD and poorer executive function in bvFTD), these differences were not absolute. Rather, there was graded variation with considerable overlap along these dimensions (Fig. 2). The phenotypic overlap occurred alongside radiological overlap; bvFTD and SD patients did not divide absolutely along a frontal vs. temporal division. These findings are in keeping with the increasing evidence for many overlapping clinical features across FTD syndromes,^8,9,90,108–110^ meaning that although the classical syndromes clearly exist, there is considerable variation within each of them and the boundaries between them are fuzzy.

The cognitive and neuroanatomical variation in FTD can be captured by a transdiagnostic approach, whereby FTD is conceptualised as a multidimensional space in which patients represent different phenotypical points along various dimensions.^9,90,108,111^. There are two key advantages of this conceptualisation of FTD. First, a transdiagnostic approach can not only accommodate but also explain “mixed” cases who may not fall neatly into a category,^112^ and as such may be excluded from research studies/clinical trials, despite being relative common. Second, recent large-scale studies have utilised a transdiagnostic approach and applied data-driven analyses to reveal the shared clinical, cognitive and behavioural dimensions in FTD and their neurobiological mechanisms.^1,9,90,108,113,114^ This has key implications for the development of symptomatic treatments, which could target specific cognitive/behavioural dimensions that span across FTD syndromes (and potentially other neurological disorders) and stratify patients for symptomatic trials based on the presence/absence of a dimension regardless of the diagnostic label or neuropathology. Furthermore, it may be possible to titrate interventions based on an individual patient’s position across these dimensions.

### Limitations and future directions

Neuronal loss occurs relatively late in the cascade of pathology in neurodegenerative disorders.^115^ Consequently, structural MRI can be insensitive to other markers of neuropathology such as hypometabolism,^50^ synaptic loss^116^ and neurotransmitter alterations.^117^ Combining structural MRI with additional neuroimaging measures may thus provide important further insight into the neural architecture of social-semantic knowledge.

Semantic memory relies on a network of brain regions, including the bilateral ATL hub and modality-specific spokes which dynamically interact with the hub to support coherent conceptual representations.^21,22,118^ Illuminating the specific cortical “spokes” that are important for the formation of social concepts is an important topic for future research. There is ongoing interest in the role of the OFC in socially-relevant concepts, with suggestions that this region ‘tags’ social concepts with hedonic value.^11–13,119^ Evidence from neuropsychology, rTMS and computational models has shown that selective lesions/perturbations to cortical ‘spoke’ regions can generate category-specific semantic impairments,^120–122^ raising the intriguing possibility that OFC damage could selectively impair comprehension of social concepts. The widespread correlated atrophy in FTD means that disentangling category-selective deficits from a generalised semantic impairment is difficult, however future studies could explore selective social-semantic deficits in people with OFC lesions.

There is evidence from fMRI and TMS that whilst the ventrolateral ATL is crucial for all concepts including social concepts, the superior ATL also contributes but more selectively for social-semantic knowledge.^15,32,33,56,57^ One possible argument is that the lack of hemispheric differences detected in the current study occurred because the ATL atrophy in SD is centred on the anterior ventral and polar temporal aspects, with the superior ATL less consistently affected. However, the atrophy in FTD is not restricted to the inferior temporal gyrus but extends to the remaining ATL gyri (as displayed in Fig. 1).^1,123^ This distribution of atrophy means it is not possible to disentangle the differential functions of ventral versus superior ATL regions in the studies of SD. Future studies could use fMRI to explore the importance of different ATL subregions for multiple types of social concepts.

We and others have proposed that at least some of the changed behaviours associated with FTD might result from a degradation of social-semantic knowledge, in keeping with other theories of behavioural change in FTD.^13,14,16,124^ It is currently not known which specific concepts are critical to supporting social behaviours in FTD, and whether distinct behavioural profiles result from degraded conceptual knowledge from ATL atrophy vs. atrophy in other areas including the OFC, anterior cingulate cortex and insula. Future studies should formally investigate how degraded social-semantic knowledge is related to the behavioural changes in FTD and distinct from disinhibition as the cause of ‘impulsive’ challenging behaviours.

## Data availability

Due to the limits of the ethics approval for these patient studies, the data cannot be openly shared. Requests for suitably anonymised data can be addressed to the senior author and may require a data transfer agreement. Code for the analyses from this study have been made available at : https://github.com/MRouse90/Social-Semantics-FTD.

## Acknowledgements

We thank the patients and their families or caregivers for giving up the time to take part in the study. We also thank Dr. Thomas Cope, Prof. Masud Husain, Dr. Sian Thompson and Dr. Sofia Toniolo for their help with recruitment.

