## Supplementary material for "Social-semantic knowledge in frontotemporal dementia and after anterior temporal lobe resection"

### SUPPLEMENTARY METHODS

#### General semantic and background neuropsychology tasks

##### Modified camel and cactus test

The camel and cactus test (CCT) is a 64-item task and assesses the ability to detect semantic associations (for example, matching 'owl' with 'bat' because they are both nocturnal animals), and forms part of the Cambridge Semantic Memory Test Battery.[1,2] This study used the recently developed modified-CCT, which comprises 32 items and uses photographs as items as opposed to a combination of photographs and line drawings.[3] For each item, participants are shown a photo at the top of a screen, and four photos underneath, and are instructed to point to the photo goes best with the photo at the top.

##### Cambridge Naming task

The Cambridge Naming task is part of the Cambridge Semantic Memory Test Battery.[1] Participants are shown a line drawing,[4] and asked to name them. Half of the items are living (animals and fruit), and half are non-living (household items, tools and vehicles). The original version has 64-items, but a shortened 32-item version was used in the current study to minimise testing burden.[5]

##### Synonym judgement task

Participants are shown a word at the top of the screen, and required to point to one of three words below which goes best with the word at the top meaning (e.g., matching 'river with 'stream' and not with 'doctor').[6] The task includes both concrete and abstract words. The original version has 96-items,[6] however this study used the reduced 48-item version.[5]

##### Boston Naming Test

The Boston Naming Test is more difficult than the Cambridge Naming task, and thus was included as an additional naming task designed to be more sensitive to milder anomia.[7]

Participant are shown line-drawings, rank ordered by difficulty and asked to name them. A reduced 30-item version was used to reduce testing burden.[8]

#### Brixton Spatial Anticipation Test

The Brixton Spatial Anticipation Test is neuropsychological test of executive function.[9] The test assesses the ability to detect and follow a visuospatial rule. Participants are shown a page displaying ten circles where one is coloured blue. The location of the blue circle changes on each page, and participants must pick up on the pattern and indicate the predicted location of the blue circle on the next page. The pattern changes without warning, and participants are required to detect and learn the new pattern. The total number of errors (maximum = 55) is summed, and a scaled score derived ranging from 1-10 (1 = impaired, 2 = abnormal, 3 = poor, 4 = low average, 5 = moderate average, 6 = average, 7 = high average, 8 = good, 9 = superior, 10 = very superior).

#### Raven's Progressive Matrices

The Raven's progressive matrices is a common measure of executive function and assesses non-verbal reasoning.[10] The Coloured Progressive Matrices Set B was used in the current study, developed for mentally impaired individuals. Participants are shown an 2x2 matrix pattern of shapes, with one shape missing, and must identify the shape that completes the pattern out of six possible response options.

### UK-English version of the Social Norms Questionnaire

Instructions: Following is a list of behaviours that a person might engage in. Please decide whether or not it would be socially acceptable and appropriate to do these things in the mainstream culture of the UK and answer yes or no to each. Think about these questions as if they were occurring in front of or with a stranger or acquaintance, NOT a close friend or family member.

**Would it be socially acceptable to:**

1. Tell a stranger you don't like their hairstyle?
2. Spit on the floor?
3. Blow your nose in public?
4. Ask a coworker their age?
5. Cry during a film at the cinema?
6. Push in a queue if you are in a hurry?
7. Laugh when you yourself trip up and fall?
8. Eat pasta with your fingers?
9. Tell a coworker your age?
10. Tell someone your opinion of a film they haven't seen?
11. Laugh when someone else trips and falls?
12. Wear the same shirt everyday?
13. Keep money you found on the pavement?
14. Pick your nose in public?
15. Tell a coworker you think they are overweight?

16. Eat ribs with your fingers?
17. Tell a stranger you like their hairstyle?
18. Wear the same shirt twice in two weeks?
19. Tell someone the ending of a film they haven’t seen?
20. Hug a stranger without asking first?
21. Talk out loud during a film at the cinema?
22. Tell a coworker you think they have lost weight?

### SUPPLEMENTARY TABLES

**Supplementary Table 1 Voxel-based morphometry results**

| | Hemisphere | Number of voxels | Peak MNI co-ordinate | | | Peak MNI co-ordinate | *t*-value |
|---|---|---|---|---|---|---|---|
| | | | *x* | *y* | *z* | | |
| *FTD < Controls* | | | | | | | |
| | Bilateral | 263,655 | -27 | 9 | -21 | Temporal pole | 13.79 |
| | Right | 3,352 | 29 | -71 | -50 | Cerebellum | 5.06 |
| | Left | 1,570 | -47 | -74 | -50 | Cerebellum | 4.02 |
| *bvFTD < Controls* | | | | | | | |
| | Bilateral | 147,625 | 2 | 36 | -15 | Gyrus rectus | 8.94 |
| *SD < Controls* | | | | | | | |
| | Bilateral | 227,548 | -30 | -5 | -42 | Inferior temporal gyrus | 21.82 |
| | Right | 3,582 | 29 | -69 | -48 | Cerebellum | 4.80 |
| *Left TLE < Controls* | | | | | | | |
| | Left | 48,006 | -29 | -20 | -17 | Hippocampus | 24.63 |
| *Right TLE < Controls* | | | | | | | |
| | Right | 51,989 | 26 | -14 | -20 | Hippocampus | 29.26 |
| *SD < bvFTD* | | | | | | | |
| | Left | 10,071 | -18 | 6 | -39 | Temporal pole | 6.35 |
| | Right | 2,607 | 15 | -6 | -38 | Undefined | 5.53 |

bvFTD = behavioural-variant frontotemporal dementia, FTD = frontotemporal dementia, MNI = Montreal Neurological Institute, TLE = temporal lobe epilepsy, SD = semantic dementia

**Supplementary Table 2 Percentage of FTD participants given chance-level score on each task**

| Test | bvFTD | SD |
|---|---|---|
| ACE-R Total | 0 | 0 |
| MMSE | 0 | 0 |
| ACE-R Attention | 0 | 0 |
| ACE-R Memory | 0 | 0 |
| ACE-R Fluency | 0 | 0 |
| ACE-R Language | 0 | 0 |
| ACE-R Visuospatial | 0 | 0 |
| Cambridge Naming | 0 | 0 |
| Boston Naming | 0 | 0 |
| Camel and cactus test | 11.5 | 13.6 |
| Synonym judgement | 15.4 | 18.2 |
| Brixton | 15.4 | 9.1 |
| Raven's | 7.7 | 9.1 |
| Face-name matching | 11.5 | 13.6 |
| Face-profession matching | 15.4 | 18.2 |
| Landmark-name matching | 11.5 | 13.6 |
| Famous Face matching | 15.4 | 13.6 |
| Unfamiliar face matching | 15.4 | 13.6 |
| Abstract social synonym judgement | 15.4 | 13.6 |
| Abstract non-social synonym judgement | 15.4 | 13.6 |
| Social word-picture matching | 7.7 | 4.5 |
| Non-social word-picture matching | 7.7 | 4.5 |
| Basic emotion matching | 15.4 | 9.1 |
| Complex emotion matching | 15.4 | 9.1 |
| Social Norms Questionnaire | 7.7 | 18.2 |
| TASIT-Sarcasm | 15.4 | 18.2 |

ACE-R = Addenbrookes Cognitive Examination-Revised, bvFTD = behavioural-variant frontotemporal dementia, MMSE = Mini Mental State Examination, SD = semantic dementia, TASIT = The Awareness of Social Inference Test

**Supplementary Table 3 Regions of grey matter intensity associated with factor scores**

| | Hemisphere | Number of voxels | Peak MNI co-ordinate | | | Peak MNI co-ordinate region | t-value |
|---|---|---|---|---|---|---|---|
| | | | x | y | z | | |
| *FTD Severity* | | | | | | | |
| | Left | 33,707 | -42 | 6 | 36 | Precentral gyrus | 6.37 |
| | Right | 3,488 | 30 | 26 | 54 | Superior frontal gyrus | 6.54 |
| *Semantic memory* | | | | | | | |
| | Left | 26,501 | -44 | 5 | -15 | Temporal pole | 6.77 |
| | Right | 19,843 | 45 | 26 | -30 | Temporal pole | 6.68 |

FTD = frontotemporal dementia, MNI = Montreal Neurological Institute

### SUPPLEMENTARY FIGURES

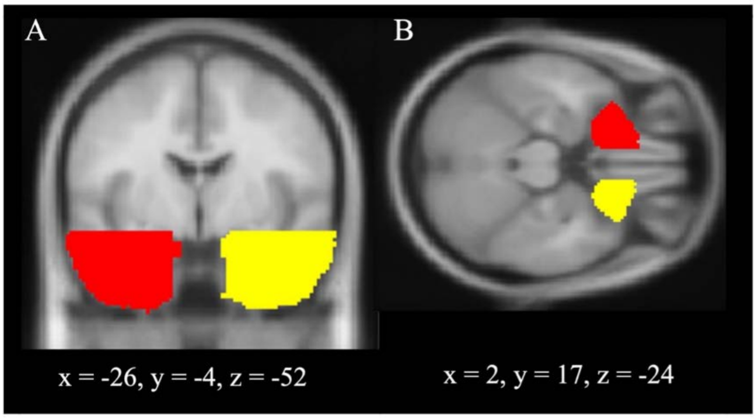


**Supplementary Figure 1. Frontotemporal indices.** Locations of the regions of interest used to calculate magnitude and asymmetry indices. **(A)** The left anterior temporal lobe ROI (shaded in red) and the right anterior temporal lobe ROI (shaded in yellow). **(B)** The left orbitofrontal cortex ROI (shaded in red) and the right orbitofrontal cortex ROI (shaded in yellow). Co-ordinates are reported in Montreal Neurological Institute space.

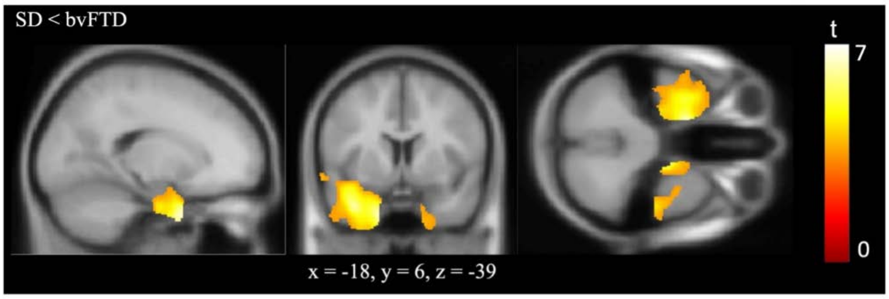


**Supplementary Figure 2. Grey matter differences between FTD subtypes.** Regions of reduced grey matter volume in SD compared to bvFTD. Groups were compared using independent t-tests, with age, intracranial volume and scanner site included as covariates. Images are thresholded using a cluster-level threshold of $Q < 0.05$ after an initial voxel-level threshold of $P < 0.001$. Significant clusters are overlaid on the MNI avg152 T1 template. Co-ordinates are reported in Montreal Neurological Institute space.

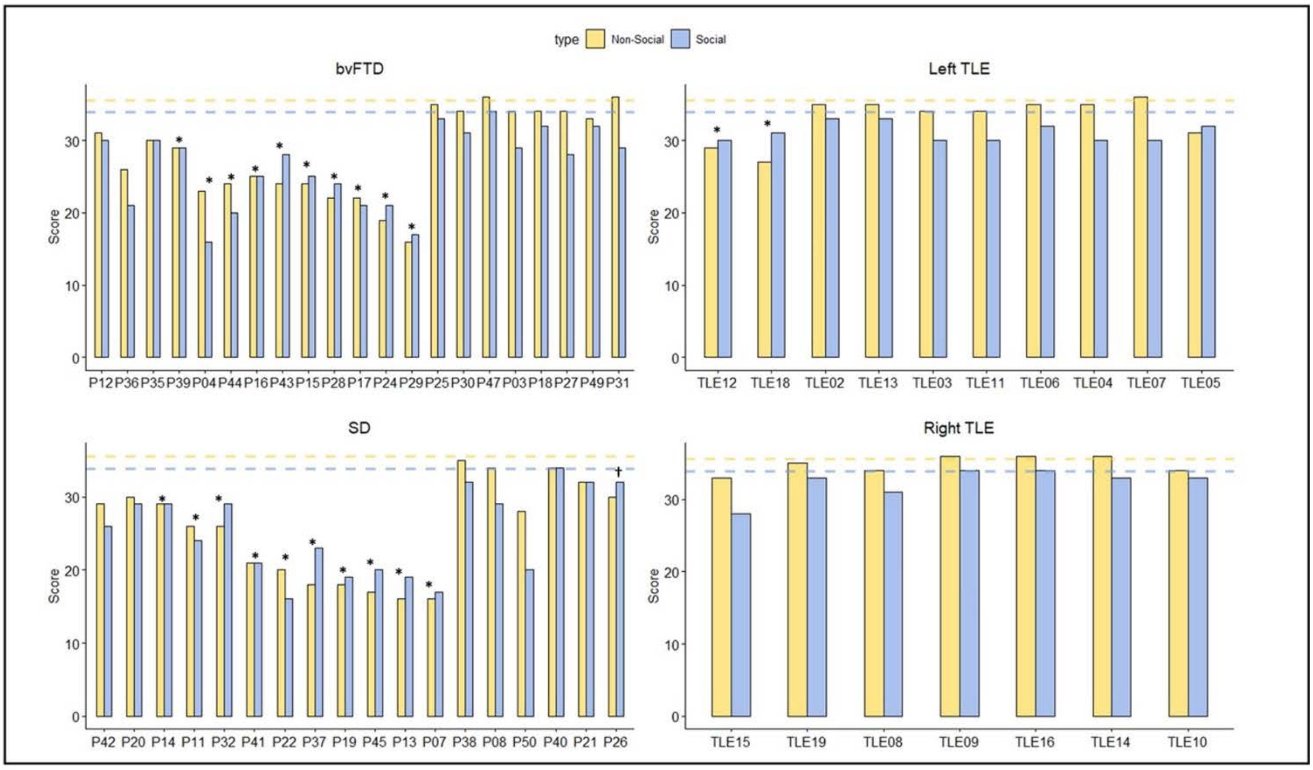


**Supplementary Figure 3. Performance on the abstract social and abstract non-social synonym judgement tasks.** Dashed horizontal lines indicate the control average on each task. ***Strong dissociation †Classical dissociation

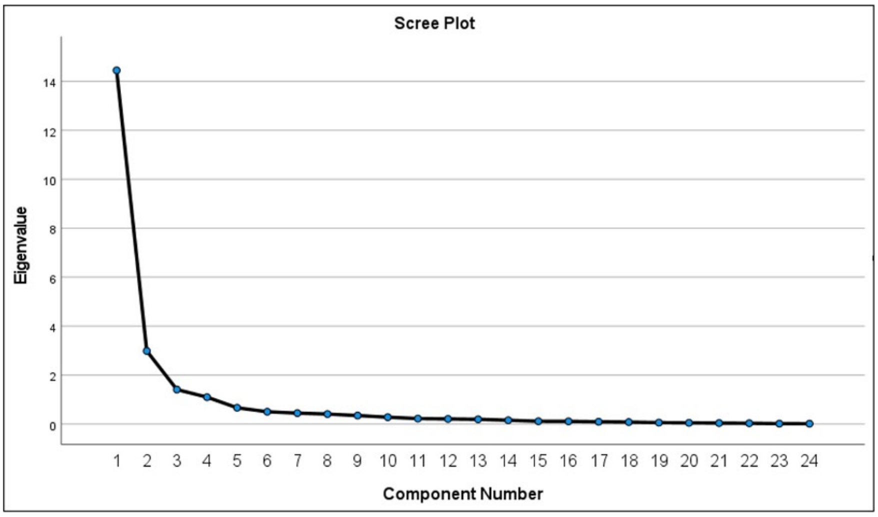


**Supplementary Figure 4. Scree plot displaying eigenvalues for each principal component.**

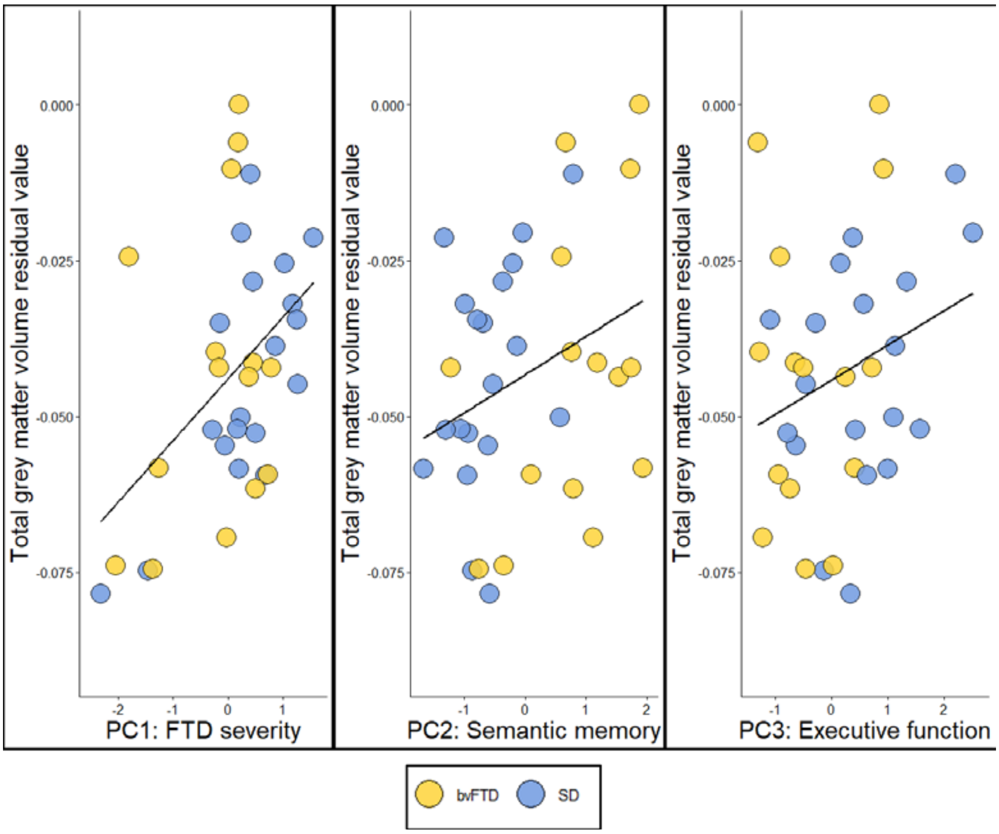


**Supplementary Figure 5. Association between total grey matter volume and factor scores in FTD.** Each data point represents the total grey matter volume index (residual values based on linear models fitted using the control data) plotted against (A) *FTD severity* factor scores, (B) *Semantic memory* factor scores and (C) *Executive function* factor scores.
